# Modeling Vaccine Failures and Behavioral Change: Effects on Disease Transmission Dynamics and Thresholds

**DOI:** 10.1101/2025.02.21.25322689

**Authors:** Irasema Pedroza-Meza, M. Adrian Acuña-Zegarra, Jorge X. Velasco-Hernández

**Affiliations:** Departamento de Matematicas, Universidad de Sonora, Boulevard Luis Encinas y Rosales s/n, Col. Centro, Hermosillo, 83000, Sonora, Mexico; Instituto de Matemáticas, UNAM-Juriquilla, Blvd. Juriquilla No. 3001, Juriquilla, 76230, Queretaro, México

**Keywords:** Mathematical model, Vaccine efficacy, Imperfect vaccines, Backward bifurcation, Kermack-McKendrick

## Abstract

Vaccination is a cornerstone of infectious disease control, yet vaccines are not fully protective, leaving a fraction of the vaccinated population susceptible to infection. This partial protection can alter behavior, as individuals who perceive themselves as immune may reduce adherence to preventive measures. Motivated by this, we investigate how behavioral changes among non-immune vaccinated individuals influence the dynamics of a directly transmitted disease and the basic reproduction number. We propose a model that incorporates vaccine failure through three facets (take, degree, and duration) alongside a behavioral parameter that modifies contact rates according to compliance with mitigation measures.

Our analysis highlights the critical role of the behavioral index in key phenomena, including backward bifurcation and overall disease dynamics. We identify two thresholds. The first specifies the values of the behavioral index for which backward bifurcation does not arise, thereby indicating the conditions under which the disease may persist. The second establishes a relationship between the behavioral index and vaccine efficacy, which allows us to compare the transmission dynamics of our model with those of the classical vaccination model.

## 1. Introduction

Motivated by the absence of complete immunity from vaccines against respiratory diseases and the behavioral changes induced by non-pharmaceutical interventions [1, 2, 3, 4, 5], this study explores the impact of the behavior of non-immune vaccinated individuals on disease transmission dynamics. For this aim, we formulate an extension of classic epidemiological models [6], incorporating vaccination. Following [7], we assume that the vaccine may exhibit three facets of vaccine failure: “take”, “degree”, and “duration.” We model these failures by (i) splitting the vaccinated population into two classes (immune vaccinated and non-immune vaccinated), (ii) incorporating a reduction in the transmission contact rate for the non-immune vaccinated population, and (iii) assuming that immune vaccinated individuals become fully susceptible after a specific period of time. In addition, we explore whether the proposed model presents similar phenomena to those observed in classical models that include vaccination such as backward bifurcation. These modeling considerations motivate a closer examination of how vaccine efficacy and behavioral responses jointly influence epidemic outcomes.

In particular, understanding the interplay between vaccine efficacy and behavioral changes can provide insight into the effectiveness of control and mitigation strategies. The effects and efficiency of control and mitigation strategies are one of the central interests in the study of the spread of epidemics [8, 9, 10, 11, 12, 13]. Vaccines are a fundamental tool for the control and eradication of infectious diseases. However, the beneficial effects of the vaccine on a population can be diminished by behaviors that increase the contact rate. Vaccine efficacy is a measure of how well a vaccine protects against infection, hospitalization, and death [14]. Vaccine efficacy is not full; there is always a certain percentage of vaccinated individuals for whom the vaccine does not confer protection or the protection is weaker than the expected average [15, 16, 17]. Risk-averse and risk-prone behaviors play a significant role in the outcome of the effectiveness of a vaccine. Risk-averse behaviors reduce the contact rate, strengthening the vaccine’s efficacy; on the contrary, risk-prone behaviors may increase the per capita contact rate, thus increasing the average transmissibility of the pathogen. When the last occurs, vaccinated individuals, who have weak protection, increase the susceptible pool of individuals available for disease infection, which may lead, at the population level, to the existence of multiple endemic equilibria [e.g., 18, 19, 20, 21, 22].

On the other hand, studying the consequences of different vaccine properties, particularly efficacy, on the dynamics of an epidemic outbreak is closely associated with the conditions under which oscillatory dynamics or multiple equilibrium points occur in epidemiological differential equation systems [see e.g., 18, 23, 24]. Usually, multiple endemic equilibria, resulting from a backward bifurcation, appear when the basic reproduction number is below 1 [25, 26, 18, 27, 28, 29]. Therefore, one of our objectives in this work is to study the behavioral change in generating this atypical subthreshold phenomenon.

Many mathematical models assume that imperfect immunization reduces the effective contact rate [see e.g., 30, 31, 12, 32]. From a modeling perspective, we characterize a vaccine using three essential parameters. The first is coverage, the percentage of the population vaccinated. The second is waning immunity, defined as the period during which a vaccinated individual remains protected before losing the immunity conferred by the vaccine. The third is vaccine efficacy, the proportion of vaccinated individuals who are genuinely protected [14]. We refer to the combination of these three parameters—coverage, efficacy, and waning time—as the vaccination policy, with the occurrence of a backward bifurcation depending on their magnitudes [33].

It is important to note that classical models often define vaccine efficacy solely in terms of protective efficacy. This definition captures only the reduction in disease risk, not the fraction of individuals who develop immunity and are therefore fully protected from infection (effective immunity of the vaccine). Despite this distinction, reported efficacy figures generally follow the classical definition of protective efficacy. In the recent COVID-19 pandemic, several vaccines achieved efficacies above 90% [34, 35], while others ranged between 70% and 80% [36, 37]. For other infectious diseases, reported efficacies vary widely [38, 39]. In this study, we model both processes separately.

In what follows, we present and analyze a mathematical model that incorporates behavioral change and vaccine failures. We formulate our mathematical model in Section 2. Section 3 presents the qualitative analysis,including the existence of a backward bifurcation. Section 4 presents simulations exploring the impact of the behavioral index on disease transmission dynamics and the basic reproductive number. Finally, we draw our conclusions in Section 5.

## 2. Mathematical model

The classic models incorporating vaccination assume that the susceptible population is vaccinated at a specific rate and that vaccine efficacy is reflected in the reduction of the contact rate: the higher the efficacy of the vaccine, the greater the percentage reduction in contact rate [18, 33, 31]. This usual assumption may lead to a backward bifurcation. Backward bifurcation is a transcritical bifurcation in which the branch of endemic equilibria arises with a negative slope, allowing for the existence of multiple endemic states below the threshold value of the reproductive number. In this case, the system exhibits two locally asymptotically stable states and one unstable equilibrium point.

One of the simplest models that addressing this phenomenon is based on the one proposed by [18]

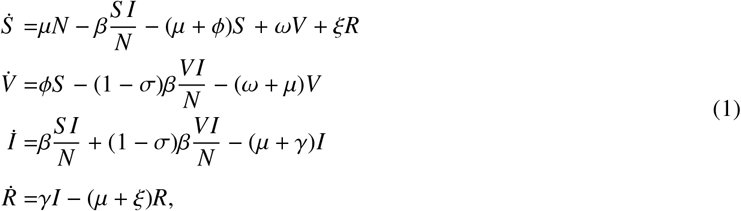

where *S*, *V, I*, and *R* denote the susceptible, vaccinated, infected, and recovered populations, respectively. The vaccine-induced immunity rate is represented by *ω*, vaccination coverage represents the number of individuals vaccinated per unit of time (*ϕS*), and *σ* can also be interpreted as the protective efficacy, i.e., *σ* is the proportional reduction in the contact rate, so that 1 − *σ* represents the degree of vaccine failure. Backward bifurcation can appear for specific ranges of (*ϕ, ω, σ*) [18].

This study extends the classic vaccination model by incorporating vaccine failures [7, 40]. This approach allows us to distinguish the effects of behavioral change from those arising solely from reductions in the transmission contact rate among vaccinated individuals. We represent take failure (all-or-nothing vaccines) by dividing the vaccinated population into two groups: *immune vaccinated* (*V*_+_), who acquire absolute immunity, and *non-immune vaccinated* (*V*_−_), who develop weak or no immunity. Degree failure (leaky vaccines) is a reduction (1 − *σ*) in the transmission contact rate of *V*_−_ individuals. Finally, the loss of immunity (i.e., duration failure) is incorporated into both vaccinated classes through the average waning time 1/*ω*, which represents the duration of protection before immunity wanes. In addition, we assume that vaccination status may influence adherence to non-pharmaceutical interventions (NPIs). While immunized individuals (*V*_+_) do not contribute to disease transmission, non-immunized vaccinated individuals (*V*_−_) do. The latter may decrease, maintain, or increase their transmission contact rate depending on the extent to which they adopt, maintain, or relax non-pharmaceutical interventions (NPIs); social distancing is a representative example. Some studies have shown that behavioral changes in populations, particularly during large-scale social events, can significantly alter transmission dynamics [41, 42, 43]. Therefore, it is pertinent to incorporate this effect into the dynamics of transmission. To do so, we capture this effect through a behavioral index, *ψ*, which modulates the transmission contact rate of the *V*_−_ class.

Following the ideas of [32], the vaccination rate (*ϕ*) is defined in terms of the target coverage (*p*), i.e., the fraction of the population to be vaccinated, and the time horizon *T*. Let *S* (*t*) denote the normalized fraction of unvaccinated susceptible individuals at time *t*, with initial condition *S* (0) = 1. Assuming a constant vaccination process applied to the unvaccinated population, the dynamics can be described by the decay equation

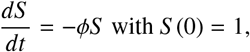

whose solution is *S* (*t*) = *e*^−*ϕt*^. Therefore, the target population fraction of vaccinated susceptible individuals at a given time horizon *T* is

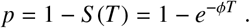

Solving for *ϕ* gives the vaccination rate:

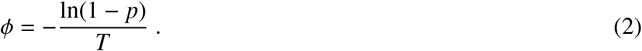

As mentioned earlier, our model incorporates take failure, which implies that the vaccinated population is split into two classes, *V*_+_ and *V*_−_. Following a similar process to derive *ϕ*, we define the rate at which susceptible individuals are successfully vaccinated as *ϕ*_+_. Let *pϵ* denote the effective vaccinated proportion, where *ϵ* represents the effective immunity of the vaccine. The effective vaccinated proportion at a given time horizon *T* is therefore 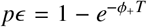. Thus, the effective vaccination rate is

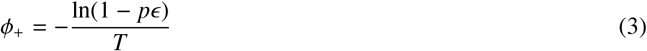

Then, assuming that *ϕ* = *ϕ*_+_ + *ϕ*_−_, where *ϕ*_−_ represents the vaccination failure rate, it follows from eqs. (2) and (3) that

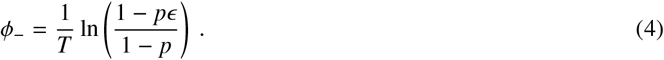

Note that *ϕ*_+_ and *ϕ*_−_ are well-defined for any *T* > 0 and can be adjusted via the target coverage *p* ∈ [0, 1). Moreover, if the effective immunity of the vaccine is null (*ϵ* = 0), then no one acquires immunity (*ϕ*_−_ = *ϕ* and *ϕ*_+_ = 0). Conversely, if the effective immunity of the vaccine is complete (*ϵ* = 1), then all vaccinated individuals acquire immunity (*ϕ*_+_ = *ϕ* and *ϕ*_−_ = 0).

### 2.1. Model equations

The structure of the proposed model, incorporating the assumptions described above, is illustrated in Figure 1.

**Figure 1.**
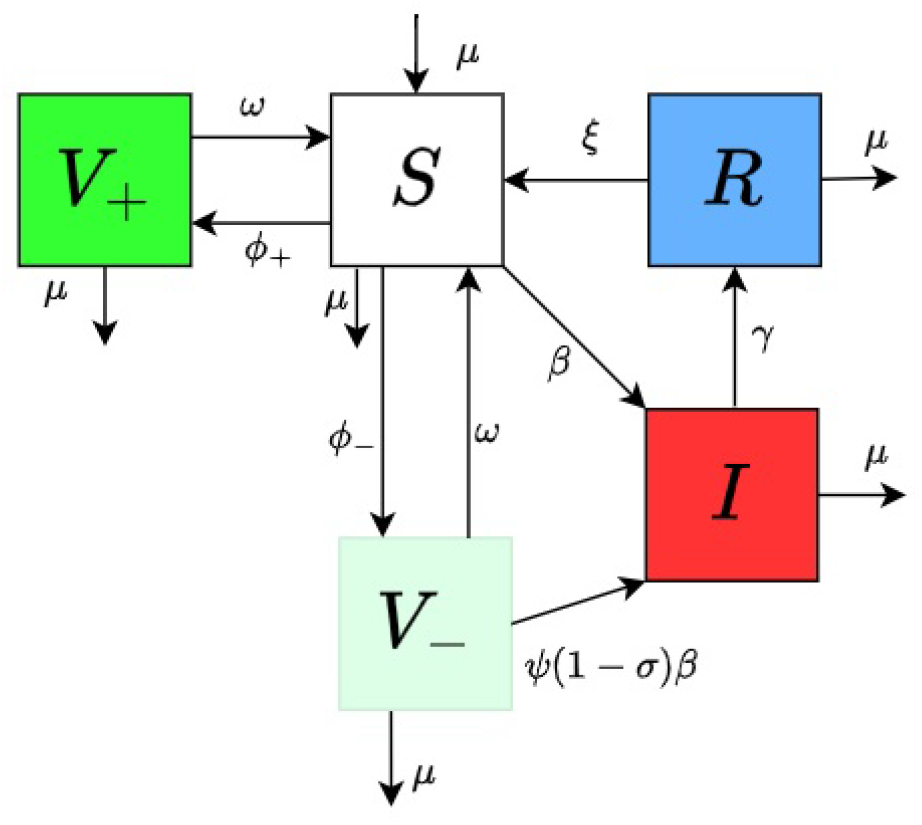
Compartmental diagram of proposed model. Here, susceptible, immune vaccinated, non-immune vaccinated, infected and recover, are represented by *S*, *V*_+_, *V*_−_, *I* and *R* respectively. Table 1 contains the definition of parameters.

Following the hypotheses previously stated, we present our mathematical model:

**Table 1:** Parameters definitions for Model (5).

| Parameter | Names |
| --- | --- |
| $\beta$ | transmission contact rate |
| $\mu$ | natural death rate |
| $\gamma$ | recovery rate |
| $1/\omega$ | waning time |
| $1/\xi$ | period of natural immunity |
| $\sigma$ | protective efficacy |
| $\psi$ | behavioral index |
| $\phi_-$ | vaccination failure rate |
| $\phi_+$ | effective vaccination rate |

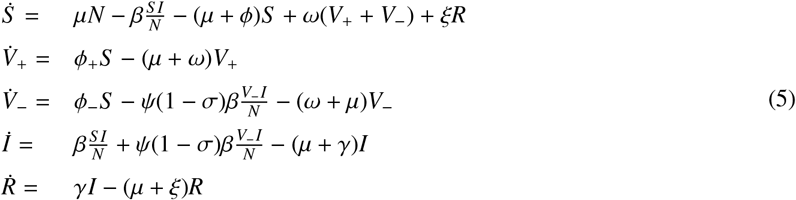

where *N* is the total population size, i.e. *N* = *S* + *V*_+_ + *V*_−_ + *I* + *R* which is constant. Table 1 summarizes parameter definitions of the Model (5).

To simplify the analysis, we rescale the total population to *N* = 1. Under this normalization, Model (5) reduces to four equations by substituting *S* = 1 − *V*_+_ − *V*_−_ − *I* − *R*. Thus, our mathematical model becomes:

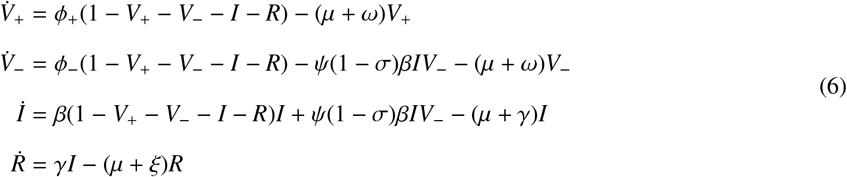

Now, we state the following proposition:

#### Proposition 1.

*Let* Ω = {(*V*_+_, *V*_−_, *I, R*)|0 ≤ *V*_+_, 0 ≤ *V*_−_, 0 ≤ *I*, 0 ≤ *R, V*_+_ + *V*_−_ + *I* + *R* ≤ 1}. *Then* Ω *is a positively invariant set under the flow of Model* (6).

Proof is provided in Appendix A.

## 3. Equilibrium states

To compute the equilibrium points, we set the right-hand sides of the Model (6) equal to zero and solve for the state variables.

### 3.1. Disease-free equilibrium and reproduction number

The disease-free equilibrium (*P*_0_) of Model (6) always exists. This equilibrium is obtained by assuming the absence of the disease (*I* = 0), rendering:

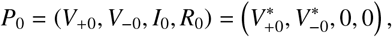

with

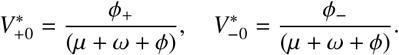

Using the method of the next generation matrix [44], we calculate the basic reproductive number (*R*_0_) of Model (6). *R*_0_ has the form

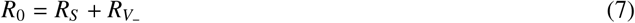

where

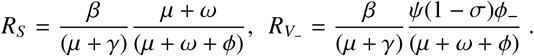

*R*_0_ can be expressed as the sum of two terms: *R*_*S*_, representing the contribution of the susceptible population to secondary infections, and 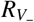, representing the contribution of the non-immune vaccinated population. The latter term also accounts for the effects of behavioral change (*ψ*), vaccination rate (*ϕ*) and protective efficacy (*σ*). In Section 4.2, we will explore the impact of *σ* on *R*_0_.

#### Proposition 2.

*The disease-free equilibrium* (*P*_0_) *is locally asymptotically stable for R*_0_ < 1 *and unstable for R*_0_ > 1.

The proof is standard and is omitted.

### 3.2. Endemic equilibria

Endemic equilibria represent states in which a disease persists in a population over time. By setting the right-hand side of Model (6) equal to zero and assuming *I* ≠ 0, the endemic equilibrium is given by the following equations:

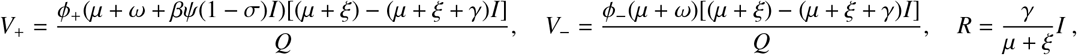

where *Q* = (*μ* + *ξ*)[(*μ* + *ω*)(*μ* + *ω* + *ϕ*) + [(*μ* + *ω* + *ϕ*_+_)*β*(*ψ*(1 − *σ*))*I*)]]. Then, *I* is obtained as the solution to:

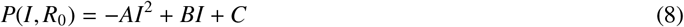

with

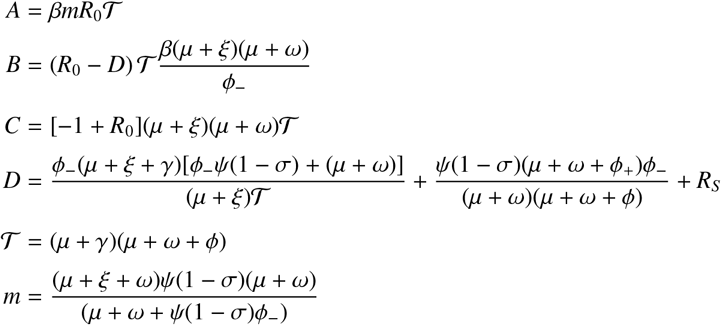

*P*(*I, R*_0_) has at most two distinct positive roots. We will show that our model exhibits a backward bifurcation under specific parameter conditions. First, we observe that if *P*(*I, R*_0_) has real positive roots, then these roots must lie within the interval (0, 1). To justify this claim, we note that *P*(*I, R*_0_) is a downward-opening quadratic polynomial in *I*. Additionally, evaluating *P*(*I, R*_0_) at *I* = 1, we obtain, for all positive *R*_0_,

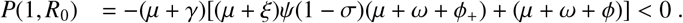

From the above, it can be concluded that if eq. (8) possesses positive roots, then these roots necessarily lie within the interval (0, 1).

Now, following [45], we demonstrate that our model exhibits a backward bifurcation under certain parameter conditions. We begin by proving the existence of a positive root in a specific case, and then we establish there is a parameter region where two positive roots coexist.

First, we set *R*_0_ = 1; then the roots of eq. (8) are

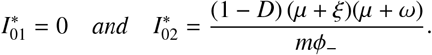

Note that 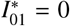 corresponds to the disease-free equilibrium (*P*_0_). On the other hand, we have that if 1 − *D* < 0, then 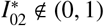, making it a biologically unfeasible root. However, if the following condition holds,

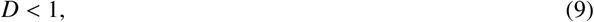

it is ensured that 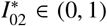, and consequently, it is a biologically feasible root. This completes the analysis for the case *R*_0_ = 1.

We now proceed to prove that eq. (8) has two positive roots. First, *P*(*I, R*_0_) is continuously differentiable function. Furthermore, we can verify that

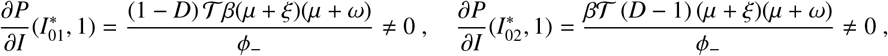

when condition (9) holds. The Implicit Function Theorem [46] guarantees the existence of a function *g*_*i*_(*R*_0_), for each of the two equilibrium points indexed by *i*, such that *P*(*g*_*i*_(*R*_0_), *R*_0_) = 0 in a neighborhood of 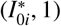. These functions represent the roots of the polynomial, which we denote by 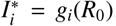. We express these functions as a series expansion around *R*_0_ = 1:

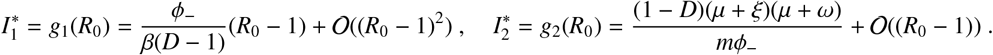

Note that, for both roots to be positive, condition (9) must hold and *R*_0_ < 1.

Next, we determine the range of *R*_0_ for which two positive roots exist. This is done by finding the positive root of eq. (8) with multiplicity two, which occurs when the discriminant of eq. (8) is equal to zero. There are two values of *R*_0_ satisfying this condition, which are:

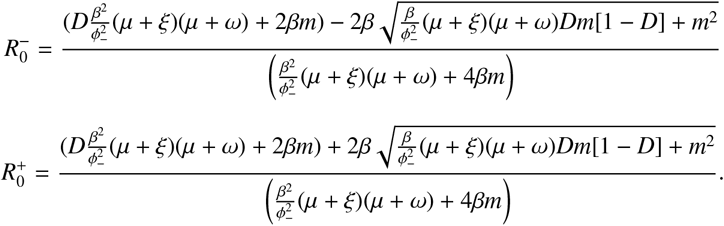

From condition (9), we deduce that 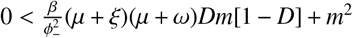. Therefore, 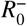 and 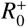 are real numbers.

Then, when 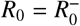, we obtain

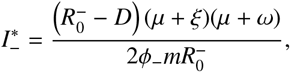

while, when 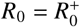, we have

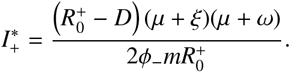

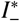 and 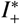 are the candidates for the positive root of eq. (8) with multiplicity two. In Appendix B, we prove that 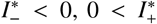, and 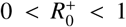. Therefore, when 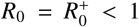, there is a unique endemic equilibrium, with multiplicity two, given by 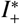. Consequently, for 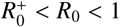, two endemic equilibria coexist. Finally, we analyze the scenario when *R*_0_ > 1. By Descartes’ Rule of Signs, eq. (8) always has a unique positive root. This implies that there is a unique endemic equilibrium.

Overall, a backward bifurcation occurs only if the contribution of susceptible individuals remains below a critical threshold, that is,

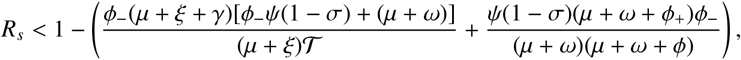

which is mainly determined by vaccination and behavioral parameters. Although these factors do not directly affect susceptible individuals, they modify the vaccinated population and, consequently, the overall reproductive number. Therefore, these parameters must be chosen appropriately to ensure that the reproductive number remains within the range permitting backward bifurcation. We formalize these results in the following proposition.

#### Proposition 3.

*Consider Model* (6) *with D, R*_0_ *and* 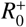 *defined above:*

a. *A unique endemic equilibrium exists if R*_0_ > 1.
b. *If D* < 1 *and* 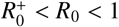 *then there are two endemic equilibria*.
c. *Otherwise, there is no endemic equilibrium*.

#### 3.2.1. Bi-stability scenario

In this section, we first illustrate the existence of a backward bifurcation (Proposition 3) and numerically explore the stability of equilibria when this phenomenon occurs. To do this, parameter values were chosen to be positive and to satisfy condition (9). Figure 2 presents the bifurcation diagram and illustrates the bistability of Model (6) when 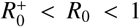. The backward bifurcation curve is generated by varying *R*_0_ as a function of behavioral index, *ψ*, (eq. (7)). To construct this curve, we assume that post-vaccination behavioral changes scale the transmission rate by a factor *ψ* ∈ [0.05, 2.9]. This interval corresponds to a reduction of up to 95% (i.e., *ψ* = 0.05) or an increase of up to 190% (i.e., *ψ* = 2.9).

**Figure 2.**
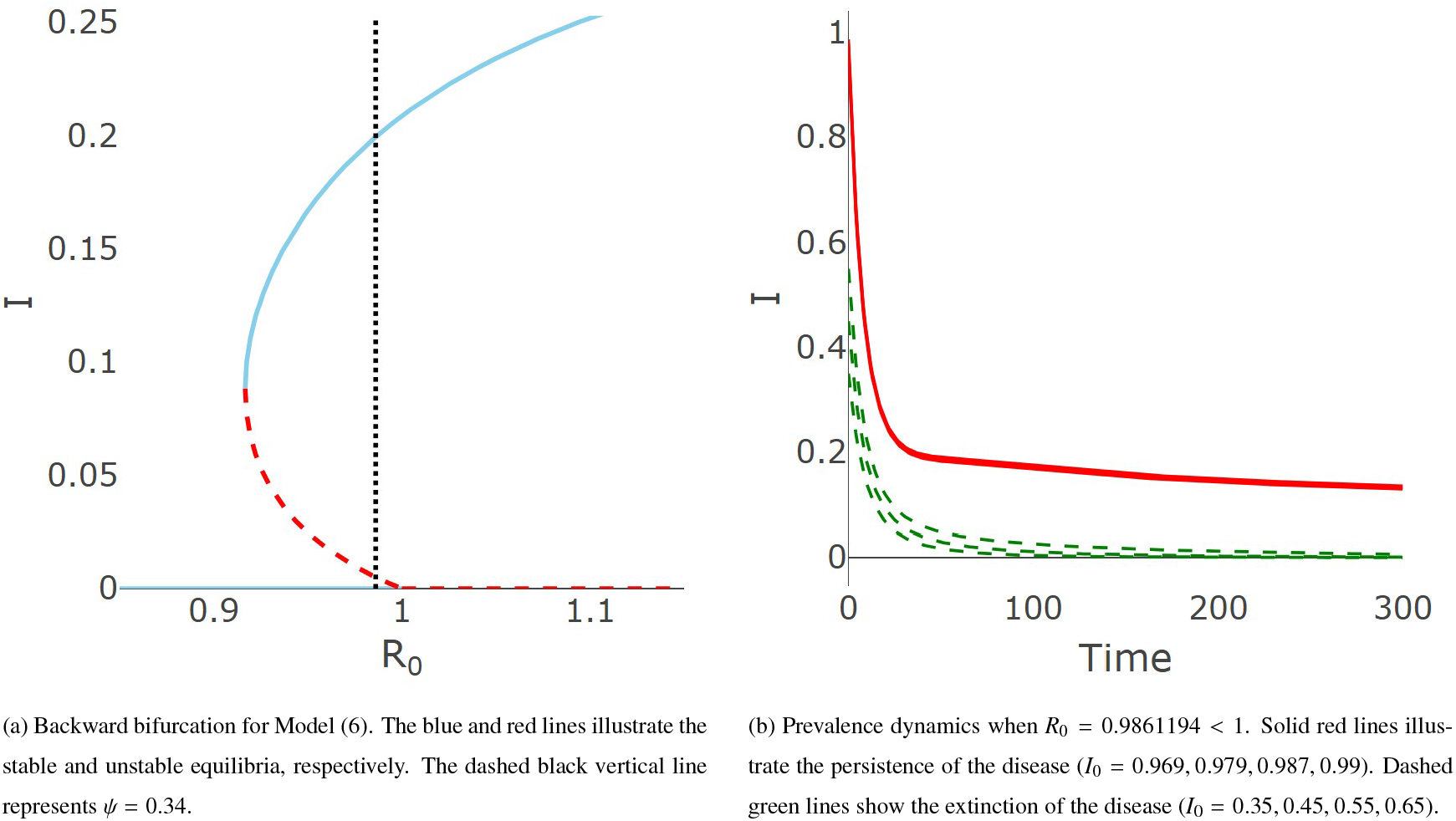
Dynamics of Model (6) when parameter values are β = 0.18001, *μ* = 0.0000391, ω = 0.002739726, γ = 0.1, ξ = 0.1, ϵ = 0.1,σ = 0.1, *p* = 0.15, and *T* = 35. (a) *ψ* varies between 0.05 and 2.9. (b) *ψ* = 0.34.

Figure 2a further illustrates the bifurcation diagram, where the solid blue line indicates stable equilibria and the dashed red line indicates unstable equilibria. This figure confirms that the disease-free equilibrium always exists, while two endemic equilibria arise when 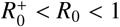. The behavior of solutions corresponding to different initial conditions is shown in Figure 2b: trajectories starting from the set highlighted in red converge to the endemic equilibrium, whereas those from the green set converge to the disease-free equilibrium. This illustrates the bistability of the system for the same set of parameter values.

Thus far, we have established that a backward bifurcation arises when the basic reproduction number is considered as the bifurcation parameter, consistent with previous studies [6, 33, 28]. To examine this phenomenon within our framework, we vary the parameter *ψ*. In Figure 3, the backward bifurcation is depicted using *ψ* as the bifurcation parameter, illustrating its relationship with the basic reproduction number for multiple values of the protective efficacy (*σ*).

**Figure 3.**
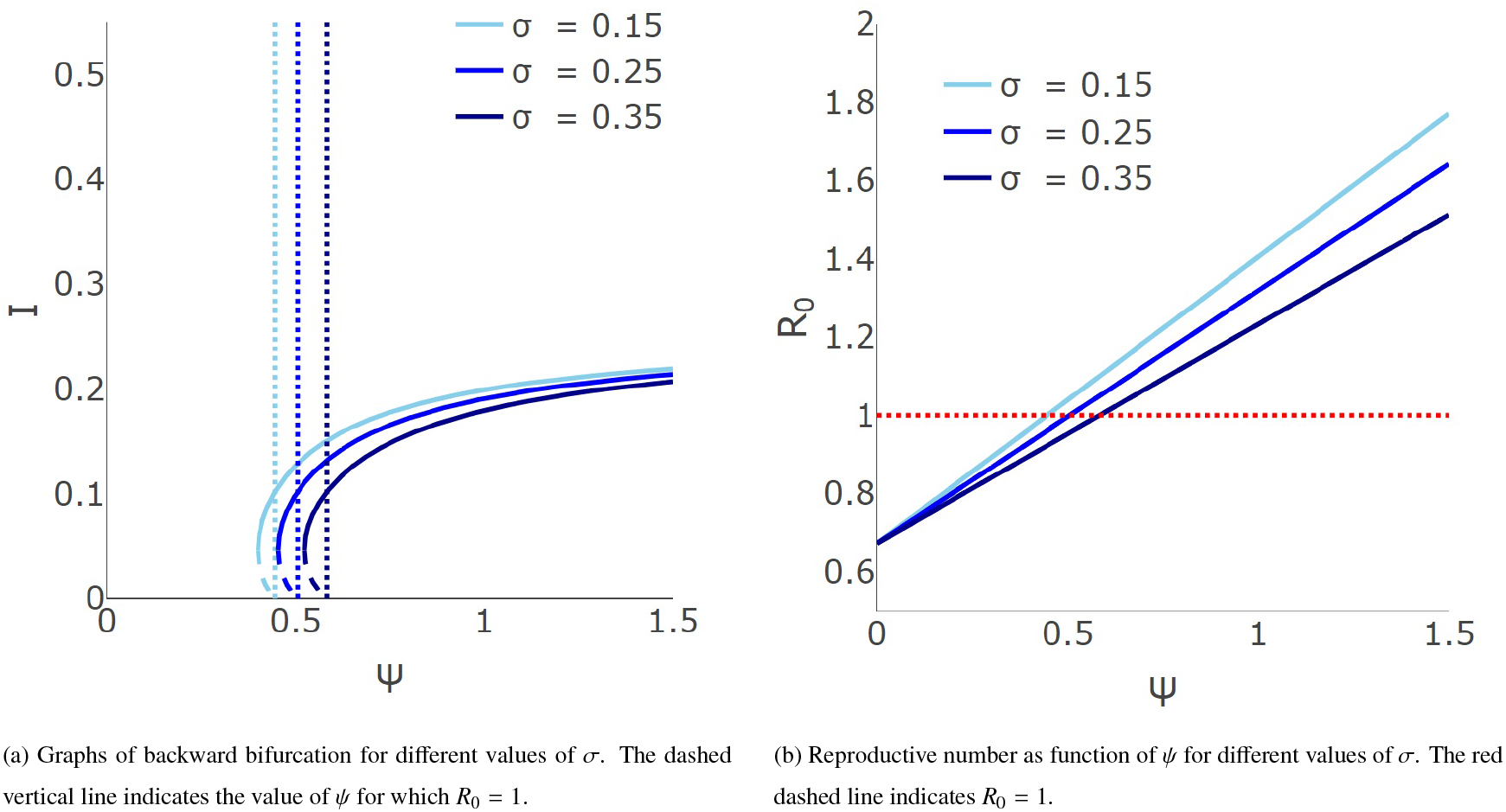
The backward bifurcation and reproductive number for different value of protective efficacy (*σ*). The parameter values used in this analysis correspond to those of Figure 2, with the exception of *ψ, σ* and *ϵ* = 0.25.

Figure 3a exhibits bifurcation diagrams for different values of *σ*, computed over the behavioral index range [0.001, 1.5] for comparability. The figure illustrates that, as protective efficacy increases, the bifurcation diagram shifts toward the right. Notably, Figure 3b illustrates the effect, observed in eq. (7), of the behavioral index *ψ* and the protective efficacy (*σ*) on the basic reproduction number (*R*_0_). Specifically, *R*_0_ is a linear function of *ψ*, and as the value of *σ* increases, the slope of this linear function decreases. Consequently, as protective efficacy increases, a greater value of *ψ* is required for condition (9) to be satisfied, and thus a backward bifurcation emerges.. As mentioned earlier, the behavioral index plays a significant role in these results; for this reason, we will examine this index in greater detail in the following section.

## 4. Role of the behavioral index (ψ)

In this section, we analyze how the behavioral index influences the dynamics of Model (6) and its basic reproduction number. We also compare the results from Models (1) and (6) under variations of *σ* and *ψ*.

Recall that the classical vaccination model does not distinguish between different types of vaccine efficacy. Thus, with the aim of making a proper comparison between Models (1) and (6), throughout this section we assume that the protective efficacy (*σ*) and the effective immunity (*ϵ*) of the vaccine, as defined in our approach, take the same values.

### 4.1. Classification of behavioral scenarios

Remember that, in this study, we model behavioral change in non-immune vaccinated individuals as a perturbation of the transmission contact rate. This behavioral change can be interpreted as the degree of compliance, maintenance, or non-compliance with mitigation measures after vaccination. Based on this interpretation, we classify this index into three categories:

a. Best case (good behavior): non-immune vaccinated individuals follow preventive measures strictly (*ψ* < 1).
b. Expected case (normal behavior): non-immune vaccinated individuals exhibit the same behavior prior to their vaccination. In this case (*ψ* = 1).
c. Worst case (bad behavior): non-immune vaccinated individuals do not comply with the mitigation measures (*ψ* > 1).

Figure 4 illustrates the effect of *ψ* on disease dynamics across the defined behavioral categories. COVID-19 parameters are employed (summarized in Table 2, with β = 0.115 except for *ψ*), and initial conditions are set to (*S* _0_, *V*_+0_, *V*_−0_, *I*_0_, *R*_0_) = (99900, 0, 0, 100, 0). In this case, for any *ψ* > 0 (with *R*_*S*_ = 0.9531008), the chosen parameter set does not satisfy condition (9) required for a backward bifurcation to occur. Blue, violet, and red lines denote “good,” “normal,” and “bad” behavior, respectively. Under “good” behavior, the disease is eradicated, whereas in the other scenarios it stabilizes at distinct endemic levels.

**Table 2:** Parameter values for Model (6) linked to COVID-19 scenario.

| Parameter | Value | References |
| --- | --- | --- |
| $\mu$ | 0.00003913 | Given<sup>a</sup> |
| $\omega$ | 0.005555556 | [47] |
| $\xi$ | 0.005555556 | [48] |
| $\gamma$ | 0.07142857 | [49] |
| $\beta$ | varying | Given<sup>b</sup> |
| $\sigma$ | 0.746 | [50] |
| $p$ | 0.5 | Given<sup>c</sup> |
| $T$ | 180 | Given<sup>c</sup> |
| $\epsilon$ | varying | Given<sup>d</sup> |

<sup>a</sup> Lifespan equal to 70 years.
<sup>b</sup> The parameter was adjusted to highlight each illustrated case.
<sup>c</sup> Target coverage of 50% to be achieved in 180 days.
<sup>d</sup> The effective immunity of the vaccine take the same values of protective efficacy.

**Figure 4.**
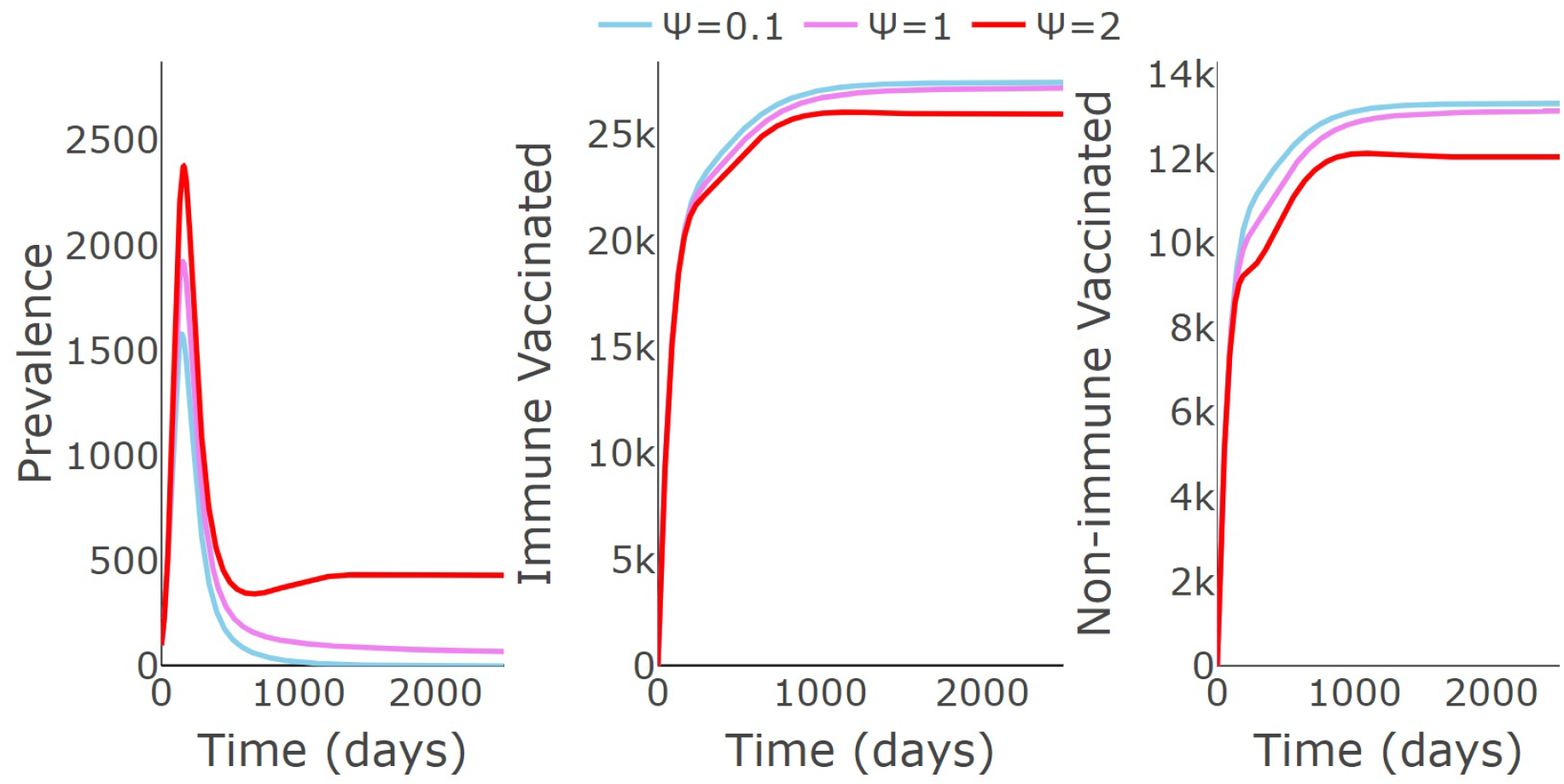
Disease dynamics under behavioral categories. Blue, violet, and red lines represent the “good” (*R*_0_ = 0.9585418), “normal” (*R*_0_ = 1.0075112), and “bad” (*R*_0_ = 1.0619216) behavioral scenarios, respectively.

#### 4.1.1. Behavioral index threshold (ψ^∗^)

In the “bad” behavior scenario, the endemic equilibrium reaches higher values compared to the other cases. This raises the question of under what circumstances ignoring mitigation and control measures makes disease eradication impossible. To address this, we introduce an expression for the behavioral index threshold. When this threshold is exceeded, the basic reproductive number is always greater than one, implying that the disease-free equilibrium will be unstable. Consider the scenario of an imperfect vaccine (σ ∈ [0, 1)), with secondary infections in the susceptible population under control (*R*_*S*_ < 1); we define the behavioral index threshold as

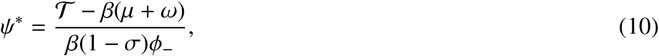

where 𝒯 = (*μ* + γ)(*μ* + *ω* + *ϕ*).

##### Proposition 4.

*Let R*_*S*_ < 1. *ψ* = *ψ*^∗^ *if and only if R*_0_ = 1.

*Proof*. We first verify that *ψ*^∗^ is biologically feasible. Since *R*_*s*_ < 1, the definition of *ψ*^∗^ (eq. (10)) ensures that *ψ*^∗^ > 0.

Then, *ψ* = *ψ*^∗^ if and only if

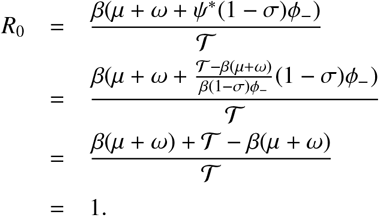

This completes the proof.

Figure 5 illustrates the impact of Proposition 4 through two scenarios: the sub-threshold regime (*ψ* < *ψ*^∗^) and the super-threshold regime (*ψ* > *ψ*^∗^). Here, β = 0.1 and the other parameter values are as in Table 2. The solid red line corresponds to the sub-threshold case, with 1.5 = *ψ* < *ψ*^∗^ = 3.618778, in which the disease disappears (*R*_0_ = 0.8997534). The dashed red line shows the super-threshold case (*ψ* = 6 > *ψ*^∗^ = 3.618778), where the disease persists (*R*_0_ = 1.1126637). It is important to note that, in both scenarios, *ψ* > 1 which corresponds to a bad-behavior regime (Section 4.1). However, even in this situation, it is still possible for the disease to die out. This suggests that individuals could adopt preventive measures more flexibly, as long as the corresponding behavioral value does not exceed the threshold. Dynamics of the other states of Model (6) are shown in the Appendix C.1.

**Figure 5.**
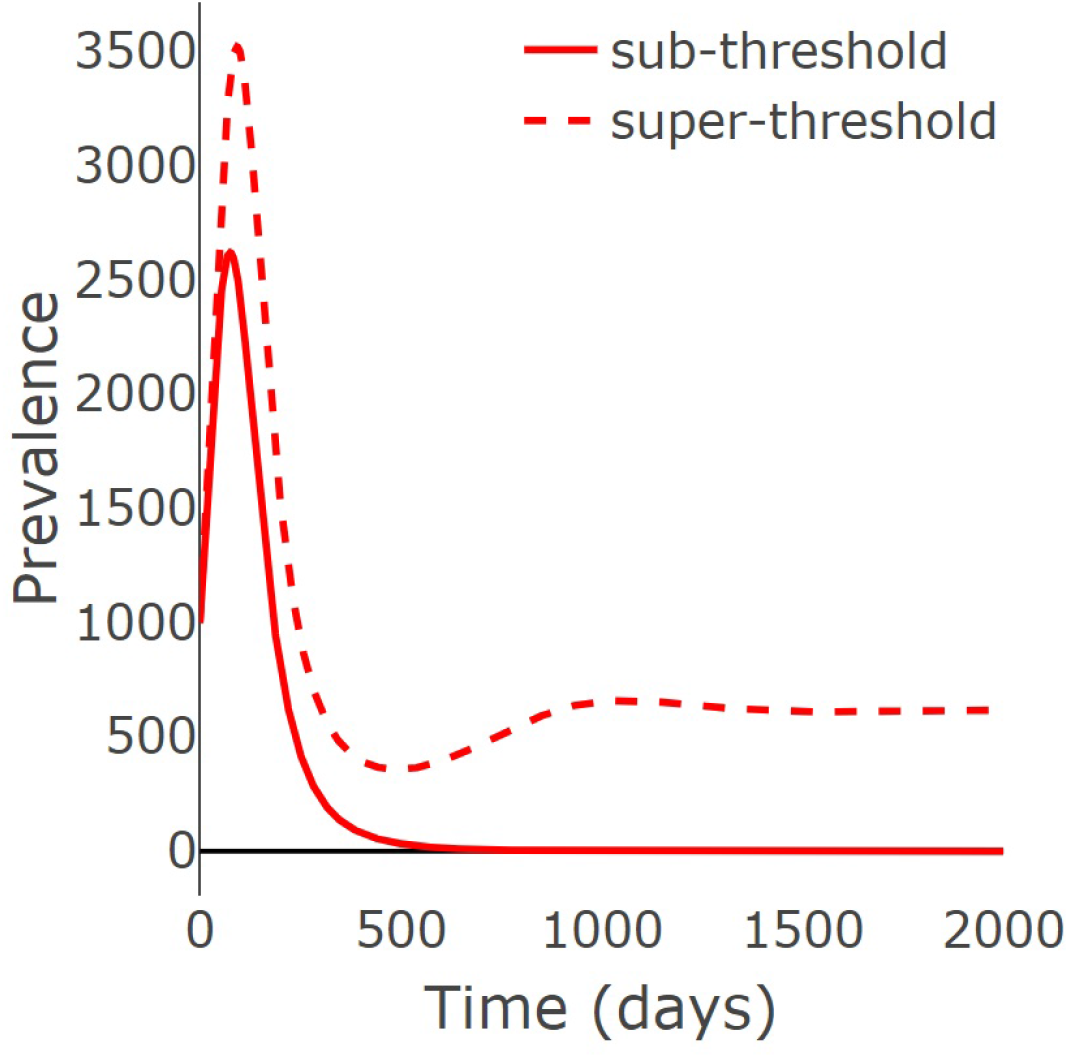
The impact of the behavioral index threshold on COVID-19 dynamics. For both cases, with initial conditions *S* (0) = 99, 000, *I*(0) = 1, 000 and *V*_+_(0), *V*_−_(0), *R*(0) = 0. We show two cases: the sub-threshold case with *ψ* = 1.5 (*R*_0_ = 0.8997534), and the super-threshold case with *ψ* = 6 (*R*_0_ = 1.1126637).

### 4.2. Comparison of Model (6) and the vaccination model (1)

In this section, we compare the results of Model (6) with those of the vaccination model (Model (1)), focusing on disease dynamics and the basic reproduction number. All parameters are as in Figure 4, except for the values of *ψ* and σ. The basic reproduction number of Model (1) is given by:

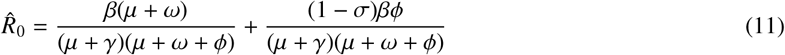

Figure 6 shows this comparison using the behavioral classification introduced in Section 4.1: “good,” “normal,” and “bad” behavior. The blue line corresponds to non-immune individuals exhibiting “good” behavior (*ψ* = 0.25), the pink line represents “normal” behavior (*ψ* = 1), and the purple line illustrates “bad” behavior (*ψ* = 3.062437). The red dashed line shows the dynamics of Model (1). Figure 6a illustrates the dynamics of the basic reproduction number as σ varies. When *ψ* ≤ 1, *R*_0_ values are consistently lower than the corresponding 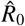 values. In contrast, for *ψ* = 3.062437, *R*_0_ exceeds 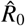 when the vaccine is not highly effective; however, as vaccine efficacy approaches 100%, the opposite trend occurs.

**Figure 6.**
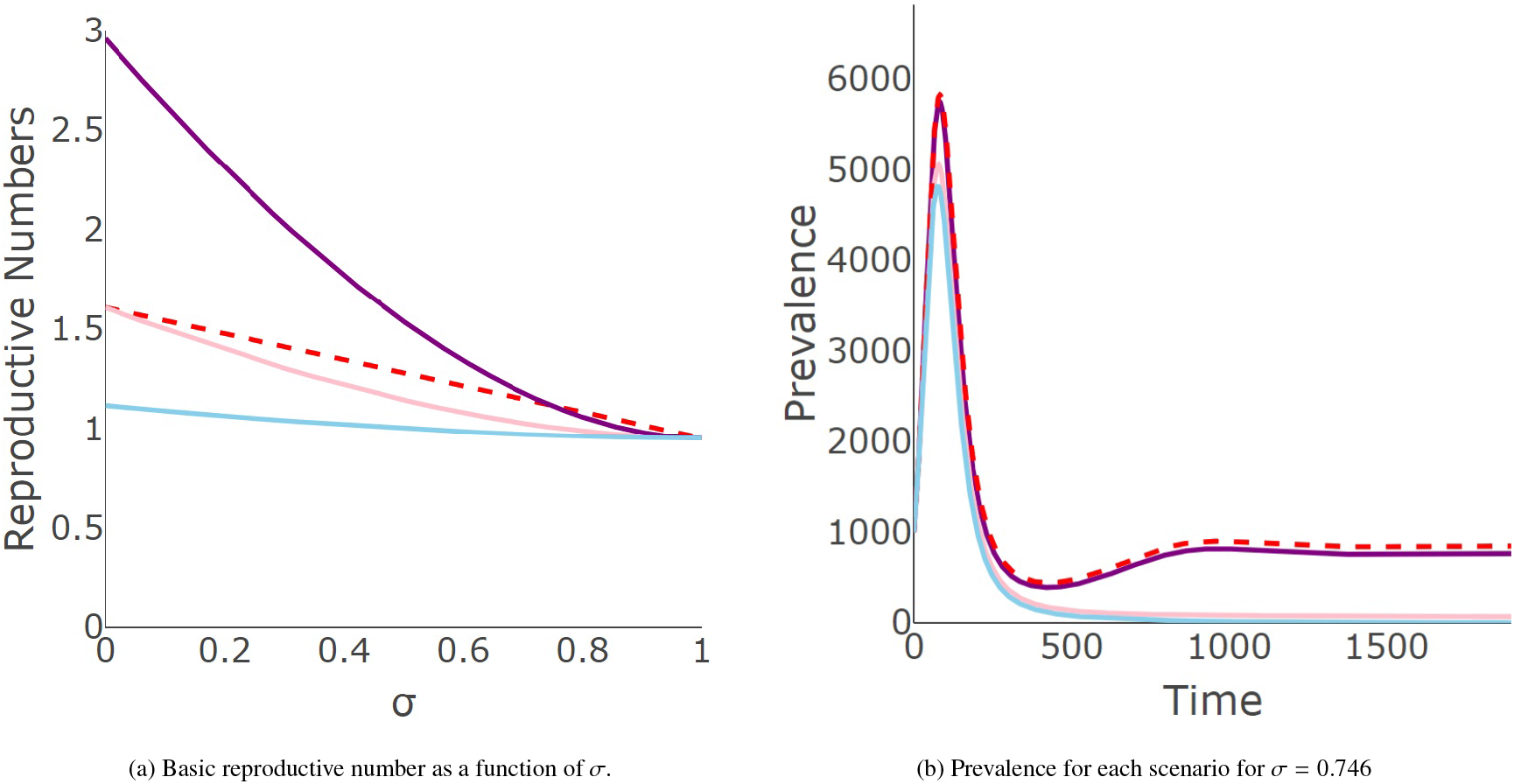
Impact of behavioral categories on Model (6) compared to Model (1). Blue, pink, and purple lines represent the dynamics of Model (6) for “good” (*ψ* = 0.25), “normal” (*ψ* = 1), and “bad” (*ψ* = 3.062437) behavior, respectively. Red dashed line illustrates dynamics of Model (1).

Figure 6b shows the prevalence for each scenario, considering the initial conditions *I* = 1000, *S* = 99000, *V*_−_ = 0, and *R* = 0. The basic reproduction numbers of Model (6) for *ψ* = 0.25, 1, and 3.062437 are 0.9667034, 1.007511, and 1.119729, respectively. For Model (1), the basic reproduction number matches that of the worst-case scenario in Model (6). Although the transient dynamic of Model (6) with “bad” behavior is similar to Model (1), the latter converges to a higher endemic level. By contrast, Model (6) scenarios with *ψ* ≤ 1 converge to a lower endemic level or the disease-free equilibrium.

Figure 6a illustrates a range of vaccine efficacy values for which Model (1) underpredicts *R*_0_, when compared to the scenario of Model (1) with *ψ* = 3.062437. Motivated by this observation, the following proposition guarantees the existence of a threshold value of the behavioral index, 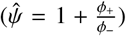, above which the basic reproduction number of Model (6) is always greater than or equal to 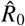.

#### Proposition 5.

*Let* ϵ ∈ [0, 1). 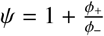, *if and only if* 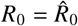.

*Proof*. Observe that 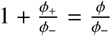. First, we suppose that 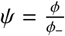, then

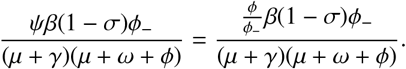

Now, we sum *R*_*S*_ in both sides,

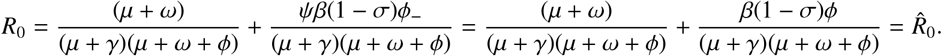

Therefore 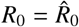. The converse implication follows directly.

The secondary threshold, 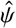, enables direct comparison between Models (1) and (6), revealing that their *R*_0_ values coincide only when non-immune individuals exhibit “bad” behavior. This threshold depends on the vaccination rate, *ϕ* = *ϕ*_+_ + *ϕ*_−_, which in turn dictates its location. Moreover, this threshold also allows Model (6) to exhibit dynamics qualitatively similar to those of Model (1) under the same set of parameters. This similarity is not limited to prevalence but can also be observed in the other states (see Appendix C.2).

## 5. Conclusions

This study examined the role of the behavioral index, *ψ*, in key phenomena, including backward bifurcation, the basic reproduction number, and overall disease dynamics. We confirm the occurrence of backward bifurcation, as reported in previous studies [18, 33, 29, 45], and explicitly highlight the role of behavior in its manifestation. Our results show that the disease-free equilibrium is stable when *ψ* is below the threshold *ψ*^∗^, and unstable when it exceeds this threshold. Moreover, Figure 3a illustrates that the behavioral index can be used as the bifurcation parameter; its critical value *ψ*^∗^ (indicated by the dashed vertical lines) plays a role analogous to the threshold value 1 when *R*_0_ is employed as the bifurcation parameter.

Furthermore, we identified an additional threshold, 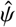, that enables the five-equation model proposed in this study to replicate the dynamics observed in the lower-dimensional model (Model (1)). Our findings highlight the interplay between the behavioral index and vaccine efficacy, showing which parameter has a greater influence on disease dynamics.

Building on the above, our results indicate that variations in population behavior may play a critical role in disease control, particularly in scenarios characterized by large outbreaks resulting from low adherence to preventive measures. For future research, it would be valuable to investigate the role of the behavioral index in other compartmental classes and to consider cases where the behavioral index dynamically depends on disease prevalence.

One limitation of this study is the inability to establish a formal relationship between backward bifurcation condition and critical thresholds. Another limitation of our work is that there is currently no data quantifying people’s adherence to preventive disease measures. Despite these limitations, our findings underscore the pivotal role of the behavioral index in capturing disease dynamics.

## Data Availability

We don’t use data

## Acknowledgments

We acknowledge support from the Joint Cooperation Fund Chile-Mexico 2022: Mathematical Modeling of Epidemic Processes, Incorporating Population Structure, Regional Distribution and Risk Groups. MAAZ and JXVH acknowledge support from the SECIHTI grant CBF-2025-G-57. We also thank Luca Ferretti and Jonathan Dushoff for their insightful comments on a previous version of this work. Finally, the authors would like to acknowledge the anonymous referees for their invaluable comments and insights on our work.

## Declarations

This work has not been published or considered for publication elsewhere. We have no conflicts of interest to disclose. We also confirm that all co-authors have agreed to the present submitted version.

## Declaration of Generative AI and AI-assisted technologies in the writing process

During the preparation of this work the authors used ChatGPT (OpenAI) in order to improve the readability and clarity of the manuscript. After using this tool, the authors reviewed and edited the content as needed and took full responsibility for the content of the published article.

## Appendix A. Invariance and positive region

### Proposition 1.

Let Ω = {(*V*_+_, *V*_−_, *I, R*)|0 ≤ *V*_+_, 0 ≤ *V*_−_, 0 ≤ *I*, 0 ≤ *R, V*_+_ + *V*_−_ + *I* + *R* ≤ 1}. Then Ω is a positively invariant set under the flow of Model (6).

*Proof*. From Model (6), we analyze the vector field on the hyperplanes where each variable is set to zero.

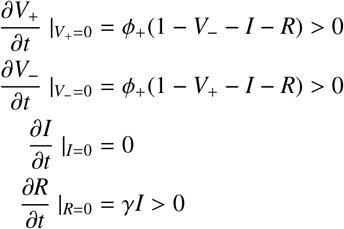

By the above, we guarantee that for an initial condition in the positive orthant, the solution remains in the positive orthant. Finally, we will prove that for any initial condition in the hyperplane 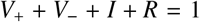, the solutions of Model (6) remain in Ω. Let 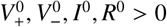, such that 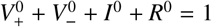, then

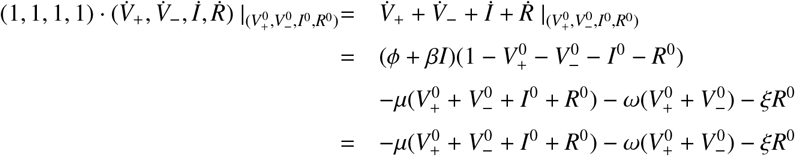

The above allows us to conclude that for any initial condition in Ω, its solution will remain in Ω.

## Appendix B. Backward bifurcation

In this section, we prove that the vertex of the parabola associated with the backward bifurcation is 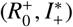.

*Proof*. We first prove that 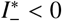. Note that, to establish this, it is sufficient to show that 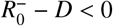, thus

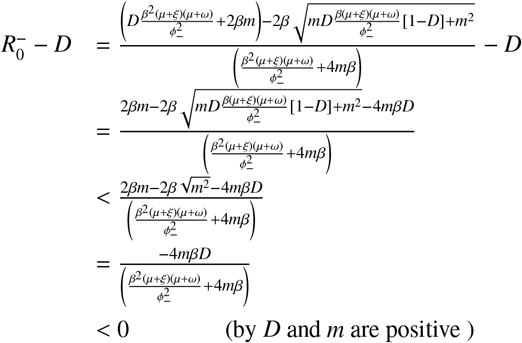

Therefore, 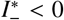. Consequently, 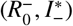 is not the vertex of the parabola associated with the backward bifurcation.

Now, we demonstrate that 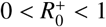. The proof that 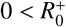 is immediate. On the other hand,

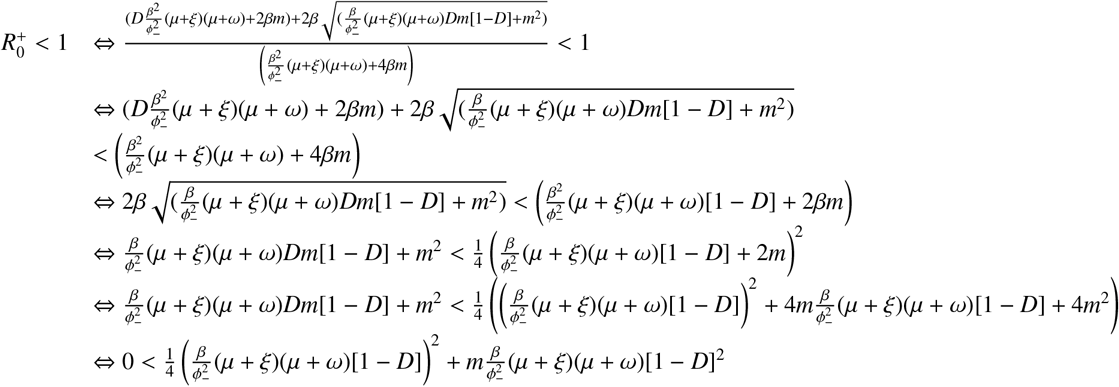

Thus, we guarantee that 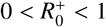. To conclude the proof, we demonstrate that 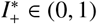. We begin by proving that 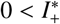, for which it is sufficient to show that 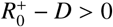. Then

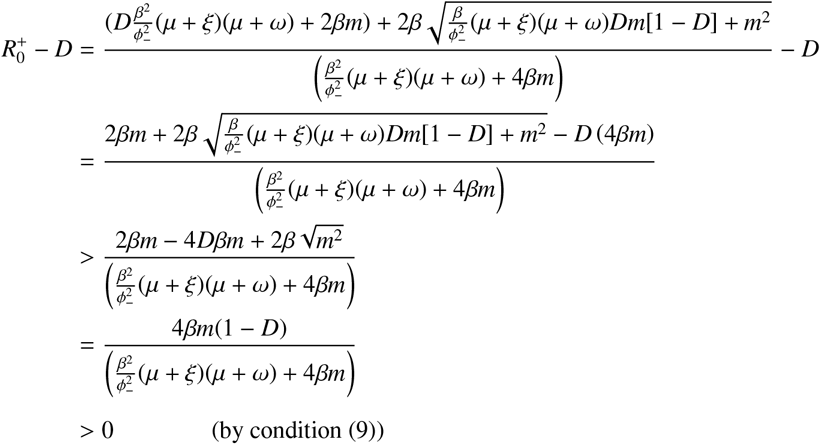

Therefore, we conclude that 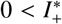. Finally, as we proved at the beginning of Section 3.2, if the root of eq. (8) is positive, then it must be less than one. Hence, 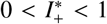. Consequently, the vertex of the parabola associated with the backward bifurcation is 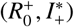.

## Appendix C. Supplementary Figures

### Appendix C.1. Complementary Figures of sub-threshold and super-threshold scenario

This section presents figures corresponding to the other populations in the sub-threshold and super-threshold scenarios, not shown in Section 4.1. Figure C.7 shows both scenarios (above and below the threshold) for susceptible and immune populations, while Figure C.8 presents the same scenarios for non-immune and recovered populations.

**Figure C.7:**
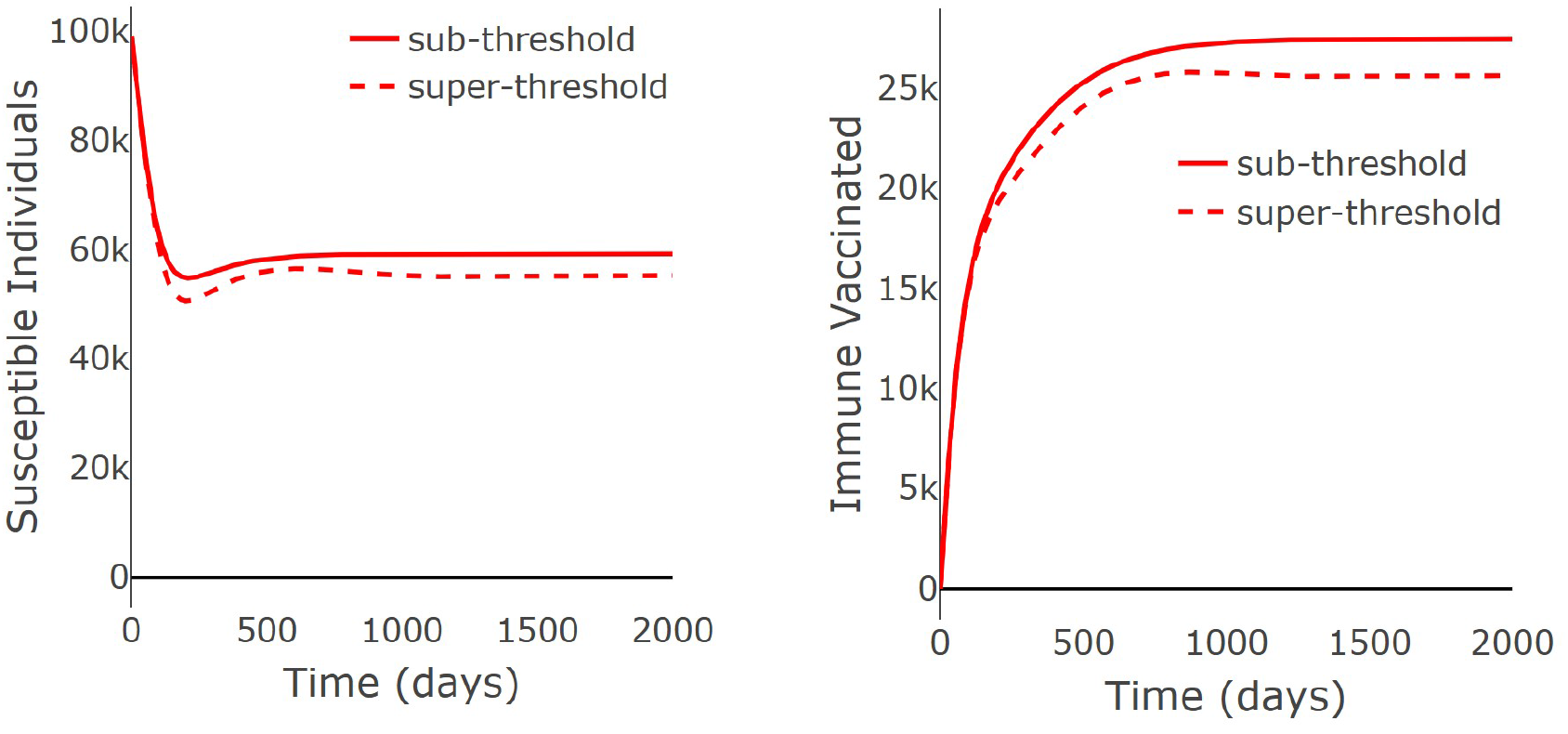
The impact of the behavioral index threshold on susceptible and immune vaccinated individuals. The parameters and the initial condition are used in Figure 5.

**Figure C.8:**
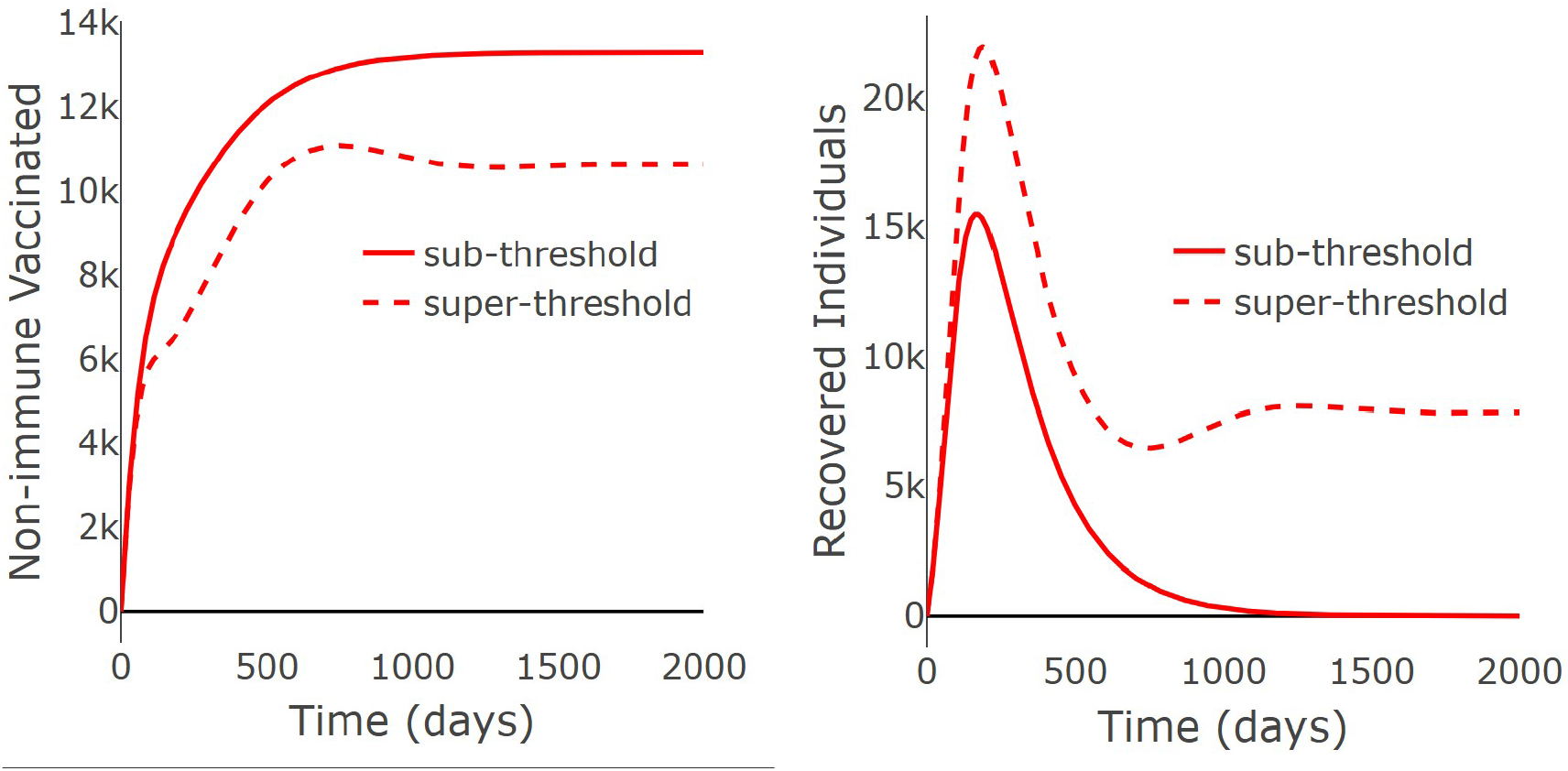
The impact of the behavioral index threshold on non-immune vaccinated and recovered individuals. The parameters and the initial condition are used in Figure 5.

#### Appendix C.2. Complementary Figures of The second threshold

This section presents figures for the populations of the system that were not included in Section 4.2. As illustrated in Figure C.9 and Figure C.10, the scenario of Model (6) qualitatively replicates the dynamics observed in the solutions of Model (6) when the behavioral index is set at its second threshold value, 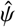.

**Figure C.9:**
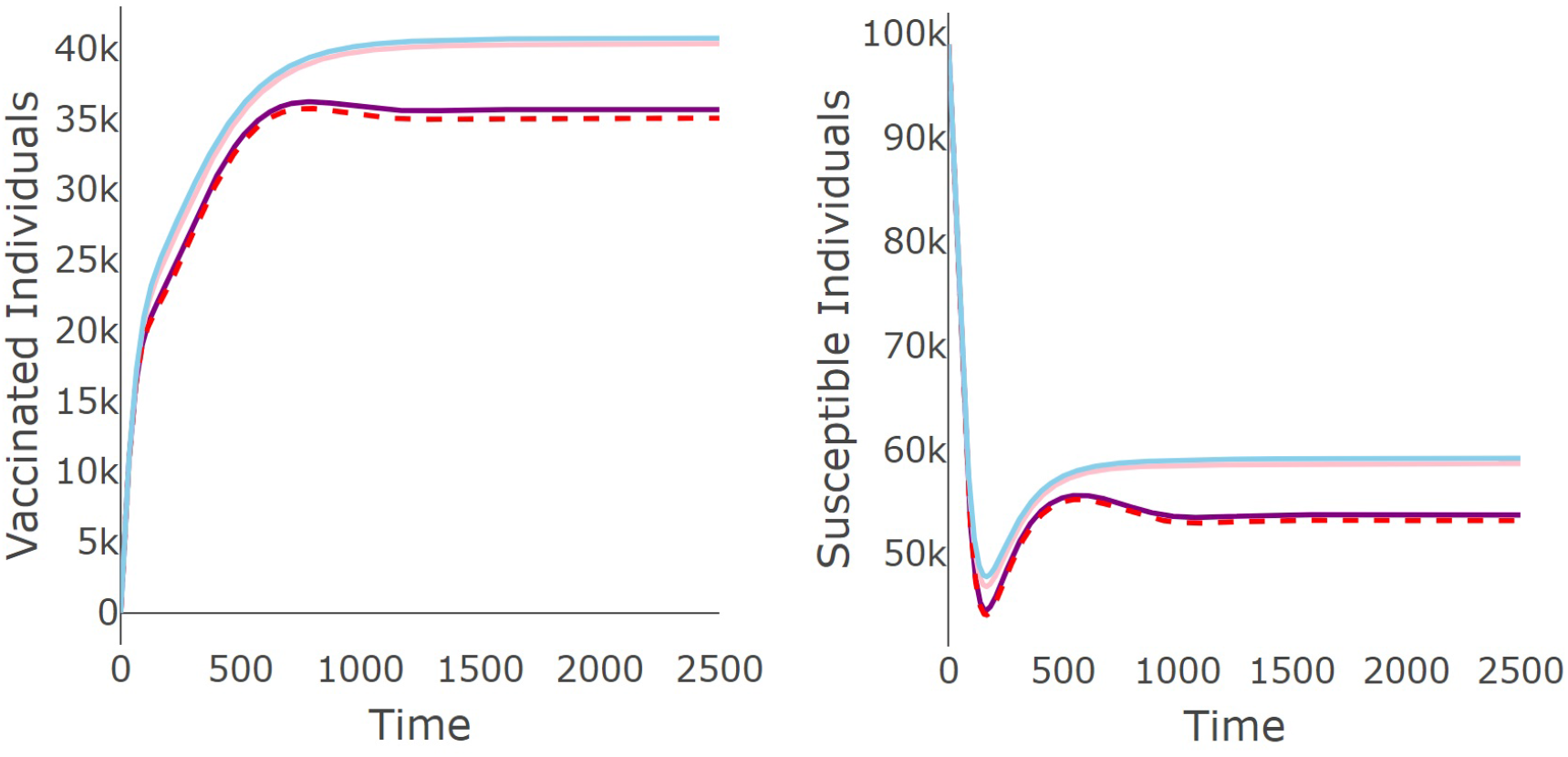
Dynamics of susceptible and vaccinated individuals for the parameters used in Figure 6b. Blue, pink, and purple lines correspond to the dynamics of Model (6) for “good,” “normal,” and “bad” behavior, respectively. Red dashed line illustrates dynamics of Model (1).

**Figure C.10:**
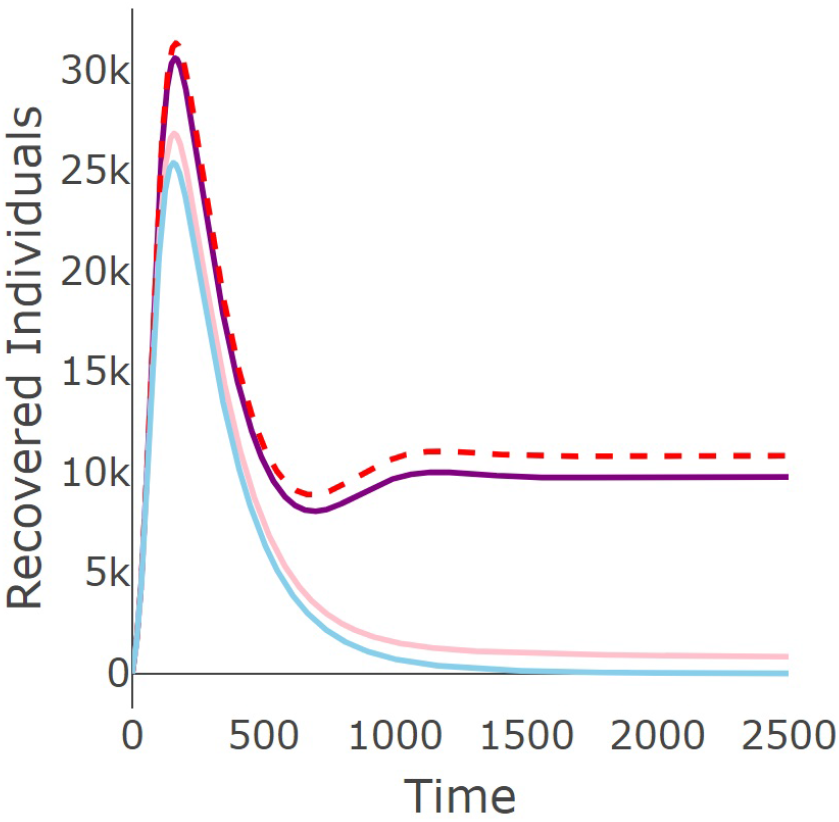
Dynamics of recovered individuals for the parameters used in Figure 6b. Blue, pink, and purple lines correspond to the dynamics of Model (6) for “good,” “normal,” and “bad” behavior, respectively. Red dashed line illustrates dynamics of Model (1).

## References

[1] H. Seale, C. E. Dyer, I. Abdi, K. M. Rahman, Y. Sun, M. O. Qureshi, A. Dowell-Day, J. Sward, M. S. Islam, Improving the impact of non-pharmaceutical interventions during COVID-19: examining the factors that in-fluence engagement and the impact on individuals, BMC Infectious Diseases 20 (2020) 1–13. doi:10.1186/s12879-020-05340-9.

[2] J. A. Lewnard, N. C. Lo, Scientific and ethical basis for social-distancing interventions against COVID-19, The Lancet infectious diseases 20 (6) (2020) 631–633. doi:10.1016/S1473-3099(20)30190-0.

[3] C. T. Bergstrom, W. P. Hanage, Human behavior and disease dynamics, Proceedings of the National Academy of Sciences 121 (1) (2024) e2317211120. doi:10.1073/pnas.2317211120.

[4] R. West, S. Michie, G. J. Rubin, R. Amlôt, Applying principles of behaviour change to reduce SARS-CoV-2 transmission, Nature human behaviour 4 (5) (2020) 451–459. doi:10.1038/s41562-020-0887-9.

[5] X. Wang, A simple proof of descartes’s rule of signs, The American Mathematical Monthly 111 (6) (2004) 525–526. doi:10.1080/00029890.2004.11920108.

[6] F. Saldaña, J.X. Velasco-Hernández, Modeling the COVID-19 pandemic: a primer and overview of mathematical epidemiology, SeMA Journal 79 (2) (2022) 225–251. doi:10.1007/s40324-021-00260-3.

[7] A. R. McLean, S. M. Blower, Imperfect vaccines and herd immunity to HIV, Proceedings of the Royal Society of London. Series B: Biological Sciences 253 (1336) (1993) 9–13. doi:10.1098/rspb.1993.0075.

[8] Y.-H. Hsieh, J. X. Velasco-Hernandez, Community treatment of HIV-1: initial stage and asymptotic dynamics, Biosystems 35 (1) (1995) 75–81. doi:10.1016/0303-2647(94)01482-M.

[9] Y.-H. Hsieh, Age groups and spread of influenza: implications for vaccination strategy, BMC infectious diseases 10 (2010) 1–12. doi:10.1186/1471-2334-10-106.

[10] V. Gemmetto, A. Barrat, C. Cattuto, Mitigation of infectious disease at school: targeted class closure vs school closure, BMC infectious diseases 14 (2014) 1–10. doi:10.1186/s12879-014-0695-9.

[11] M.A. Acuña-Zegarra, M. Santana-Cibrian, J. X. Velasco-Hernandez, Modeling behavioral change and COVID-19 containment in Mexico: A trade-off between lockdown and compliance, Mathematical biosciences 325 (2020) 108370. doi:10.1016/j.mbs.2020.108370.

[12] F. Saldaña, J.X. Velasco-Hernández, The trade-off between mobility and vaccination for COVID-19 control: A metapopulation modelling approach, Royal Society Open Science 8 (6) (2021) 202240. doi:10.1098/rsos.202240.

[13] L. Xue, X. Ren, F. Magpantay, W. Sun, H. Zhu, Optimal control of mitigation strategies for dengue virus transmission, Bulletin of Mathematical Biology 83 (2021) 1–28. doi:10.1007/s11538-020-00839-3.

[14] World Health Organization, Vaccine efficacy, effectiveness and protection (2021). URL https://www.who.int/news-room/feature-stories/detail/vaccine-efficacy-effectiveness-and-protection

[15] P. G. Smith, L. C. Rodrigues, P. Fine, Assessment of the protective efficacy of vaccines against common diseases using case-control and cohort studies, International journal of epidemiology 13 (1) (1984) 87–93. doi:10.1093/ije/13.1.87.

[16] M. E. Halloran, M. Haber, I. M. Longini Jr, C. J. Struchiner, Direct and indirect effects in vaccine efficacy and effectiveness, American journal of epidemiology 133 (4) (1991) 323–331. doi:10.1093/oxfordjournals.aje.a115884.

[17] M. E. Halloran, M. Haber, I. M. Longini Jr, Interpretation and estimation of vaccine efficacy under heterogeneity, American Journal of Epidemiology 136 (3) (1992) 328–343. doi:10.1093/oxfordjournals.aje.a116498.

[18] C. M. Kribs-Zaleta, J.X. Velasco-Hernández, A simple vaccination model with multiple endemic states, Mathematical biosciences 164 (2) (2000) 183–201. doi:10.1016/S0025-5564(00)00003-1.

[19] L. J. Allen, P. Van den Driessche, Stochastic epidemic models with a backward bifurcation, Mathematical Biosciences & Engineering 3 (3) (2006) 445–458. doi:10.3934/mbe.2006.3.445.

[20] T. K. Kar, P. K. Mondal, Global dynamics of a tuberculosis epidemic model and the influence of backward bifurcation, Journal of Mathematical Modelling and Algorithms 11 (2012) 433–459. doi:10.1007/s10852-012-9210-8.

[21] Z. Hu, W. Ma, S. Ruan, Analysis of SIR epidemic models with nonlinear incidence rate and treatment, Mathematical biosciences 238 (1) (2012) 12–20. doi:10.1016/j.mbs.2012.03.010.

[22] B. Buonomo, E. Penitente, Modelling behavioural changes and vaccination in the transmission of respiratory viruses with co–infection: B. buonomo, e. penitente, Journal of Mathematical Biology 91 (4) (2025) 41. doi: 10.1007/s00285-025-02280-3.

[23] C. M. Kribs-Zaleta, M. Martcheva, Vaccination strategies and backward bifurcation in an age-since-infection structured model, Mathematical biosciences 177 (2002) 317–332. doi:10.1016/S0025-5564(01)00099-2.

[24] J.X. Velasco-Hernández, M., A. Comas-García, D. E. N. Cherpitel, M. C. Ocampo, Superinfection between Influenza and RSV Alternating Patterns in San Luis Potosí State México, PLOS ONE 10 (3) (2015) 1–19. doi:10.1371/journal.pone.0115674.

[25] K. P. Hadeler, C. Castillo-Chávez, A core group model for disease transmission, Mathematical biosciences 128 (1-2) (1995) 41–55. doi:10.1016/0025-5564(94)00066-9.

[26] J. Dushoff, W. Huang, C. Castillo-Chavez, Backwards bifurcations and catastrophe in simple models of fatal diseases, Journal of mathematical biology 36 (1998) 227–248. doi:10.1007/s002850050099.

[27] D. Greenhalgh, M. Doyle, F. Lewis, A mathematical treatment of AIDS and condom use, Mathematical Medicine and Biology: A Journal of the IMA 18 (3) (2001) 225–262. doi:10.1093/imammb/18.3.225.

[28] J. Arino, K. Cooke, P. Van Den Driessche, J. Velasco-Hernández, An epidemiology model that includes a leaky vaccine with a general waning function, Discrete and Continuous Dynamical Systems Series B 4 (2) (2004) 479–495. doi:10.3934/dcdsb.2004.4.479.

[29] D. Benítez, I. Barradas, Panic behavior induces multiple endemic states and backward bifurcation, Mathematical Methods in the Applied Sciences 45 (5) (2022) 2831–2851. doi:10.1002/mma.7957.

[30] A. B. Gumel, Causes of backward bifurcations in some epidemiological models, Journal of Mathematical Analysis and Applications 395 (1) (2012) 355–365. doi:10.1016/j.jmaa.2012.04.077.

[31] M. Safan, F. A. Rihan, Mathematical analysis of an SIS model with imperfect vaccination and backward bifurcation, Mathematics and Computers in Simulation 96 (2014) 195–206. doi:10.1016/j.matcom.2011.07.007.

[32] M.A. Acuña-Zegarra, S. Díaz-Infante, D. Baca-Carrasco, D. Olmos-Liceaga, COVID-19 optimal vaccination policies: A modeling study on efficacy, natural and vaccine-induced immunity responses, Mathematical Biosciences 337 (2021) 108614. doi:10.1016/j.mbs.2021.108614.

[33] J. Arino, C. C. McCluskey, P. Van den Driessche, Global results for an epidemic model with vaccination that exhibits backward bifurcation, SIAM Journal on Applied Mathematics 64 (1) (2003) 260–276. doi:10.1137/S003613990241382.

[34] I. Colmegna, V. Valerio, N. Amiable, M. Useche, E. Rampakakis, L. Flamand, E. Rollet-Labelle, L. Bessette, M.-A. Fitzcharles, E. Hazel, et al., COVID-19 Vaccine in Immunosuppressed Adults with Autoimmune rheumatic Diseases (COVIAAD): safety, immunogenicity and antibody persistence at 12 months following Moderna Spike-vax primary series, RMD open 9 (4) (2023) e003400. doi:10.1136/rmdopen-2023-003400.

[35] Q. Wu, J. Tong, B. Zhang, D. Zhang, J. Chen, Y. Lei, Y. Lu, Y. Wang, L. Li, Y. Shen, et al., Real-world effectiveness of BNT162b2 against infection and severe diseases in children and adolescents, Annals of Internal Medicine 177 (2) (2024) 165–176. doi:10.7326/M23-1754.

[36] N. Ghiasi, R. Valizadeh, M. Arabsorkhi, T. S. Hoseyni, K. Esfandiari, T. Sadighpour, H. R. Jahantigh, Efficacy and side effects of Sputnik V, Sinopharm and AstraZeneca vaccines to stop COVID-19; a review and discussion, Immunopathologia Persa 7 (2) (2021) e31. doi:10.34172/ipp.2021.31.

[37] J. Sadoff, G. Gray, A. Vandebosch, V. Cárdenas, G. Shukarev, B. Grinsztejn, P. A. Goepfert, C. Truyers, I. Van Dromme, B. Spiessens, et al., Final analysis of efficacy and safety of single-dose Ad26. COV2. S, New England Journal of Medicine 386 (9) (2022) 847–860. doi:10.1056/NEJMoa2117608.

[38] C. A. Diaz Granados, A. J. Dunning, M. Kimmel, D. Kirby, J. Treanor, A. Collins, R. Pollak, J. Christoff, J. Earl, V. Landolfi, et al., Efficacy of high-dose versus standard-dose influenza vaccine in older adults, New England Journal of Medicine 371 (7) (2014) 635–645. doi:10.1056/NEJMoa1315727.

[39] R. Ray, G. Dos Santos, P. O. Buck, C. Claeys, G. Matias, B. L. Innis, R. Bekkat-Berkani, A review of the value of quadrivalent influenza vaccines and their potential contribution to influenza control, Human vaccines & immunotherapeutics 13 (7) (2017) 1640–1652. doi:10.1080/21645515.2017.1313375.

[40] F. M. G. Magpantay, M. A. Riolo, M. D. de Cellès, A. A. King, P. Rohani, Epidemiological consequences of imperfect vaccines for immunizing infections, SIAM Journal on Applied Mathematics 74 (6) (2014) 1810–1830. doi:10.1137/140956695.

[41] J. Kurland, W. E. Leal, E. M. Sorrell, N. L. Piquero, Association of National Football League Fan Attendance With County-Level COVID-19 Incidence in the 2020-2021 Season, JAMA Network Open 5 (11) (2022) e2240132–e2240132. doi:10.1001/jamanetworkopen.2022.40132.

[42] L.-C.-. R. Group, Q. Chen, M. M. Toorop, M. G. de Boer, F. R. Rosendaal, W. M. Lijfering, Why crowding matters in the time of COVID-19 pandemic?-a lesson from the carnival effect on the 2017/2018 influenza epidemic in the Netherlands, BMC Public Health 20 (1) (2020) 1516. doi:10.1186/s12889-020-09612-6.

[43] I. Dergaa, H. B. Saad, P. Zmijewski, R. Farhat, M. Romdhani, A. Souissi, J. Washif, M. Taheri, N. Guelmami, N. Souissi, et al., Large-scale sporting events during the COVID-19 pandemic: insights from the FIFA World Cup 2022 in Qatar with an analysis of patterns of COVID-19 metrics, Biology of Sport 40 (4) (2023) 1249–1258. doi:10.5114/biolsport.2023.131109.

[44] P. Van den Driessche, J. Watmough, Reproduction numbers and sub-threshold endemic equilibria for compartmental models of disease transmission, Mathematical Biosciences 180 (1) (2002) 29–48. doi:10.1016/S0025-5564(02)00108-6.

[45] J. M. Islas, R. Corona-Moreno, J.X. Velasco-Hernández, Multiple endemic equilibria in an environmentally-transmitted disease with three disease stages, Mathematical Biosciences 375 (2024) 109244. doi:10.1016/j.mbs.2024.109244.

[46] J. E. Marsden, A. J. Tromba, Vector Calculus, 6th Edition, W. H. Freeman and Company, New York, 2012.

[47] S. Diaz-Infante, M.A. Acuña-Zegarra, J.X. Velasco-Hernández, Modeling a traffic light warning system for acute respiratory infections, Applied Mathematical Modelling 121 (2023) 217–230. doi:10.1016/j.apm.2023.04.029.

[48] Y. Goldberg, M. Mandel, Y. M. Bar-On, O. Bodenheimer, L. S. Freedman, N. Ash, S. Alroy-Preis, A. Huppert, R. Milo, Protection and waning of natural and hybrid immunity to SARS-CoV-2, New England Journal of Medicine 386 (23) (2022) 2201–2212. doi:10.1056/NEJMoa2118946.

[49] R. Wölfel, V. M. Corman, W. Guggemos, M. Seilmaier, S. Zange, M. A. Müller, D. Niemeyer, T. C. Jones, P. Vollmar, C. Rothe, et al., Virological assessment of hospitalized patients with COVID-2019, Nature 581 (7809) (2020) 465–469. doi:10.1038/s41586-020-2196-x.

[50] K. R. Emary, T. Golubchik, P. K. Aley, C. V. Ariani, B. Angus, S. Bibi, B. Blane, D. Bonsall, P. Cicconi, S. Charlton, et al., Efficacy of ChAdOx1 nCoV-19 (AZD1222) vaccine against SARS-CoV-2 variant of concern 202012/01 (B. 1.1. 7): an exploratory analysis of a randomised controlled trial, The Lancet 397 (10282) (2021) 1351–1362. doi:10.1016/S0140-6736(21)00628-0.

